# Increased Risk of Alzheimer’s Disease Affected by Weight Changes but Not by Body Mass Index

**DOI:** 10.1101/2024.08.18.24312165

**Authors:** Jee Hoon Roh, Inha Jung, Hyun Jung Kim

## Abstract

**Background:** Alzheimer’s disease (AD) is an intractable and multi-factorial neurodegenerative disorder. Given the globally rapid increase in obesity and its role in AD pathogenesis, understanding the impact of body weight, its changes, and the role of physical activity on AD development can provide important guidance for preventative strategies.

**Methods:** This population-based retrospective cohort study analyzed data from Korean national health and disability databases, including 3,741,424 individuals aged 30 to 80 years at baseline, who underwent health assessments between 2003 and 2006, followed by biennial check-ups over a decade. Exposures included BMI categories (underweight, normal, overweight, obese) and body weight changes (stable, acute increase, steady increase, weight cycling, acute decrease, steady decrease). Regular physical activity was defined as consistent weekly exercise over ten years. The primary outcome was AD incidence, identified by ICD-10 codes F00 or G30. Hazard ratios (HRs) were calculated using Cox proportional hazard models adjusted for multiple risk factors.

**Results:** Baseline BMI was not significantly associated with AD incidence after adjusting for confounders, except for underweight (adjusted HR [aHR], 1.10, 95% CI, 1.05-1.15). Weight changes were significantly linked to increased AD risk, particularly weight cycling (aHR, 1.37, 95% CI, 1.35-1.40), acute decrease (aHR, 1.78, 95% CI, 1.55-2.03), and steady decrease (aHR, 1.33, 95% CI, 1.30-1.35). Regular physical activity mitigated these risks, nullifying statistical significance.

**Conclusion:** Weight changes are significant risk factors for AD, and regular physical activity mitigates these risks. Public health strategies should focus on maintaining stable weight and promoting consistent physical activity.

**Key messages:** *What is already known on this topic:* Previous studies assessing the risk of Alzheimer’s disease (AD) in relation to body weight or body mass index (BMI) have shown inconsistent results, probably due to short periods of follow-up, limited assessment of AD risk factors, and lack of washout periods to exclude the potential reverse causation between weight changes and AD development.

*What this study adds:* In an 18-year cohort study with 3741424 adults in Korea, weight changes significantly impacted the risk of AD, while baseline BMI did not. Both increases and decreases in weight, as well as weight cycling, increased the risk of AD after controlling for AD risk factors. Regular physical activity, defined as at least one exercise per week over the 10-year period, mitigated these risks.

*How this study might affect research, practice or policy:* These findings suggest that maintaining stable body weight and engaging in regular physical activity may be crucial in reducing the risk of AD, emphasizing the need for public health strategies focusing on weight stability and consistent exercise rather than simple correction of body weight.

## Introduction

Alzheimer’s disease (AD) is the most common neurodegenerative disorder among the elderly.^1^ While recent successes in monoclonal antibody trials targeting beta-amyloid shed light on disease modification, the effects have been noted only in subjects with mild cognitive impairment (MCI) and mild AD.^2^ ^3^ Thus, there remains a substantial need to modify the disease by intervening in the diverse mechanisms during its development and progression. AD has non-modifiable risk factors such as age, female sex, the presence of the apolipoprotein ε4 allele, and a family history of dementia.^4^ ^5^ Modifiable risk factors of AD include midlife obesity, reduced physical activity, midlife hypertension, diabetes mellitus (DM), hypercholesterolemia, hearing loss, and smoking.^4^ ^6^

Among the modifiable risk factors, midlife obesity is of particular interest due to its rapidly increasing prevalence worldwide^7^ and direct impact on metabolic pathways implicated in AD.^8^ ^9^ Midlife obesity not only amplifies the risk of AD but also acts as a driver for other conditions, including metabolic syndrome, which in turn may influence neurodegenerative processes.^9^ ^10^ Given this context, our study focuses on dissecting the role of body weight, its long-term changes, and potential mitigating effects of physical activity on AD development.

Results of previous studies on the risk of AD according to body weight or body mass index (BMI) are inconsistent.^11^ ^12^ Substantial research findings have indicated the effect of high body weight or BMI on AD development.^13^ A cohort study indicated a clear association between overweight, defined as a BMI over 25, and the development of AD.^14^ Another study revealed the association between higher BMI and lower brain volume in patients with AD and MCI.^15^

On the other hand, some studies indicated that being underweight in old age increased the risk of AD.^11^ ^12^ A study in subjects older than 65 years showed that weight loss may occur prior to the diagnosis of AD.^16^ Another study demonstrated that the rate of weight loss was doubled one year prior to the diagnosis of AD.^17^ A recent cohort study also reported that the risk of AD was increased in the underweight population.^16^ These findings suggest that weight loss may be an indicator of AD development. Conversely, it has also been reported that weight loss occurs during the moderate and advanced stages of AD.^18^ ^19^ Thus, whether the weight loss is a risk factor for AD, an early clinical sign of AD, or a result of behavioral changes in AD still needs to be investigated.

These conflicting results may contribute to the short duration of assessment or the diversity in demographics, including ages. Additionally, known risk factors of AD need to be considered for the interpretation of the association between AD and body weight. In addition, to remove the potential for reverse causation between body weight and AD development, an appropriate washout period is needed before tracking AD development after weight measurement. Moreover, studies assessing the impact of body weight changes on AD development are limited.

Therefore, in this 18-year cohort study in Korea, we investigated the risk of AD development according to BMI after controlling for defined AD risk factors to explore potential mechanisms underlying the conflicting results between body weight and AD. In addition, the risk of AD development was assessed by BMI changes over 10 years after the ‘post-washout period’ to refrain from the reverse causation. We also investigated the role of physical activity as a modifiable risk factor for AD development in relation to body weight, given that physical activity directly modifies obesity or body weight. We hypothesized that the risk of AD development would vary depending on the BMI or types of changes in body weight and that physical activities may affect the association.

## Materials and Methods

### Data Sources

In this population-based retrospective cohort study, data were obtained from the National Health Insurance (NHI) database, the National Health Screening Program and the National Disability Registration System. Detailed information is provided in **Supplemental Method 1**.

### Study Design and Study Subjects

The study population consisted of individuals who underwent an initial health check-up between 2003 to 2006, and then underwent 5 additional check-ups at two-year intervals over the next 10 years. Participants who missed information on birth year or BMI, and those who aged <30 or >80 years were excluded. (**Supplemental Figure 1** for the study flow diagram). We applied a one-year washout period prior to the initial check-up and a subsequent one-year post-washout following the final check-up. The index date was defined as one year after the last check-up for each participant. We excluded individuals who were diagnosed with AD prior to the index date (**Supplemental Figure 2** for the study design).

### Definitions of AD, Comorbidities, and Risk Factors

AD patients were defined as those who visited an outpatient clinic at least twice or who received inpatient treatment, under the principal diagnosis of ICD-10 code F00 (dementia in Alzheimer Disease) or G30 (Alzheimer Disease) during the monitoring period.^20^ Definitions for comorbidities and risk factors are provided in **Supplemental Method 2**.

### Categorization of BMI and Body Weight Changes

BMI (unit: kg/m^2^) was categorized into four groups according to standardized guidelines: underweight (<18.5), normal weight (18.5 to <25.0), overweight (25.0 to <30.0), or obese (≥30).^21^ Body weight change was calculated for all participants as the percent difference in weight between consecutive check-ups, over a decade with five check-ups conducted at two-year intervals. The weight (w) change rate was calculated as ((w_n_ – w_n-1_) / (w_n-1_)) × 100 and 6 weight change categories were defined: stable (-5% < weight change < +5%); acute increase (at least one increase of ≥20% without decreasing); steady increase (all increases of 5% to <20% without decreasing); weight cycling (including both increase and decrease of >5%); acute decrease (at least one decrease of ≥20% without increasing); steady decrease (all decreases of 5% to <20% without increasing).

### Assessment of Regular Physical Activity

We assessed the frequency of physical activity and participants were categorized into two groups: those who exercised consistently at least once a week over the 10-year period (regular physical activity), and those who did not (none or no regular physical activity).

#### Statistical Analyses

The study population was followed until a diagnosis of AD, death, or the end of the follow-up period on 31 December 2019. Person-years were calculated for each study subject, starting from the index date to the corresponding end date of follow-up. Study subjects contributed to person-years only when they were still at risk (i.e. alive and living in Korea without a diagnosis of AD). Incidence was calculated by dividing AD cases by the respective total person-years at risk. Incidence rates (IRs) were calculated according to BMI (underweight, normal weight, overweight, and obese) and stratified according to age and sex. Three different Cox proportional hazard regression models were used to assess the impact of BMI, body weight changes, and physical activity on AD development. Detailed methods are provided in **Supplemental Method 3**.

We used Stata version 15.0 (Stata Corp) in the execution of all statistical analyses.

The study was approved by the Institutional Review Board of the Korea University (KUIRB-2021-0328-01).

## Results

### Baseline Characteristics

Out of 4562221 participants, a total of 3741424 participants in four BMI groups (underweight: 80731; normal weight: 2420624; overweight: 1146170; obese: 93899) were finally included (**Supplemental Figure 1**). The data accounted for 20737063 person-years of follow-up. The demographics of patients distributed by baseline BMI are presented in **Table 1**. The mean age at baseline was 46.74 (±10.63) years. Approximately 65% of participants were of normal weight, while underweight and obese individuals constituted 2% and 3%, respectively.

**Table 1.**
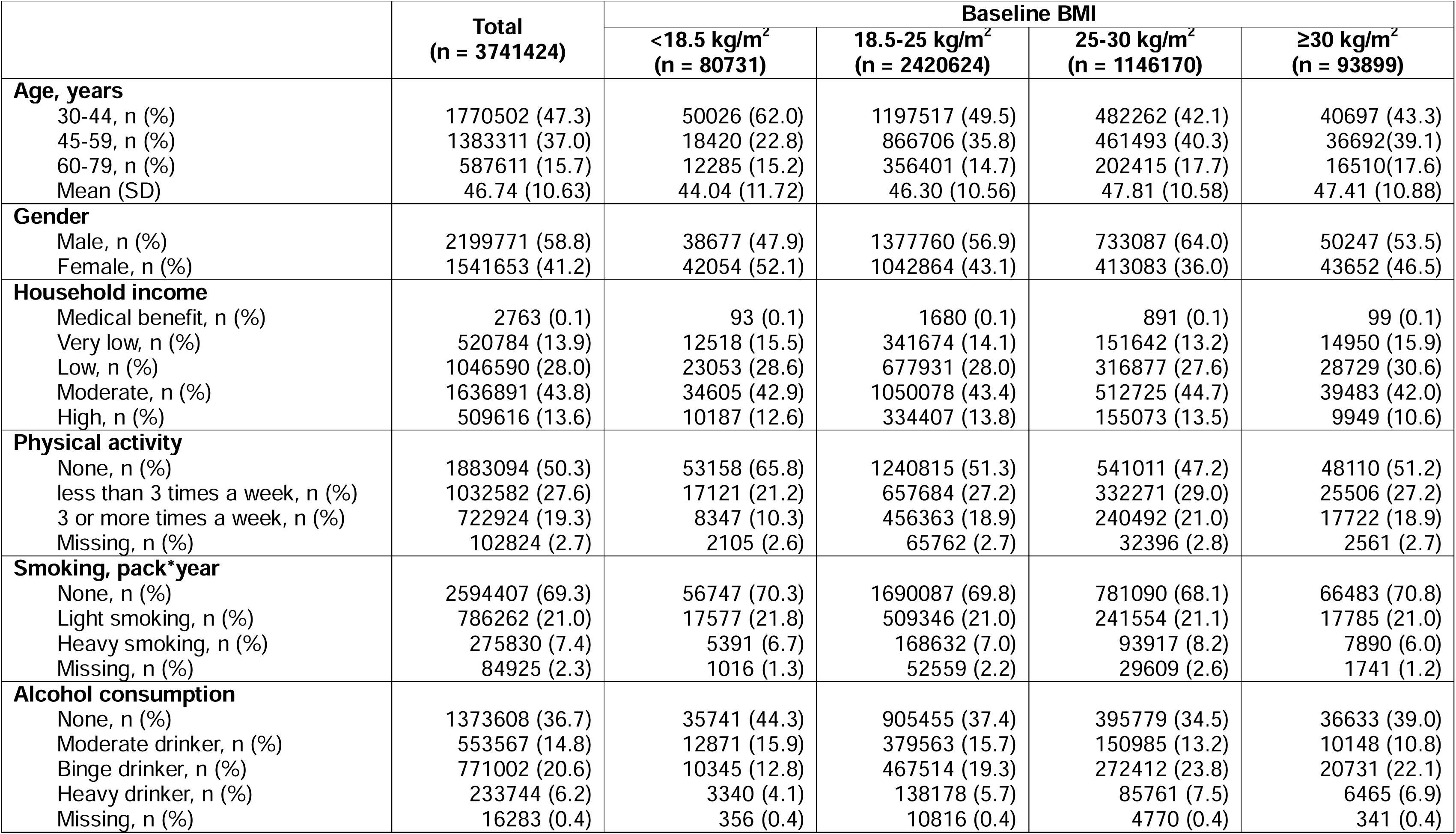
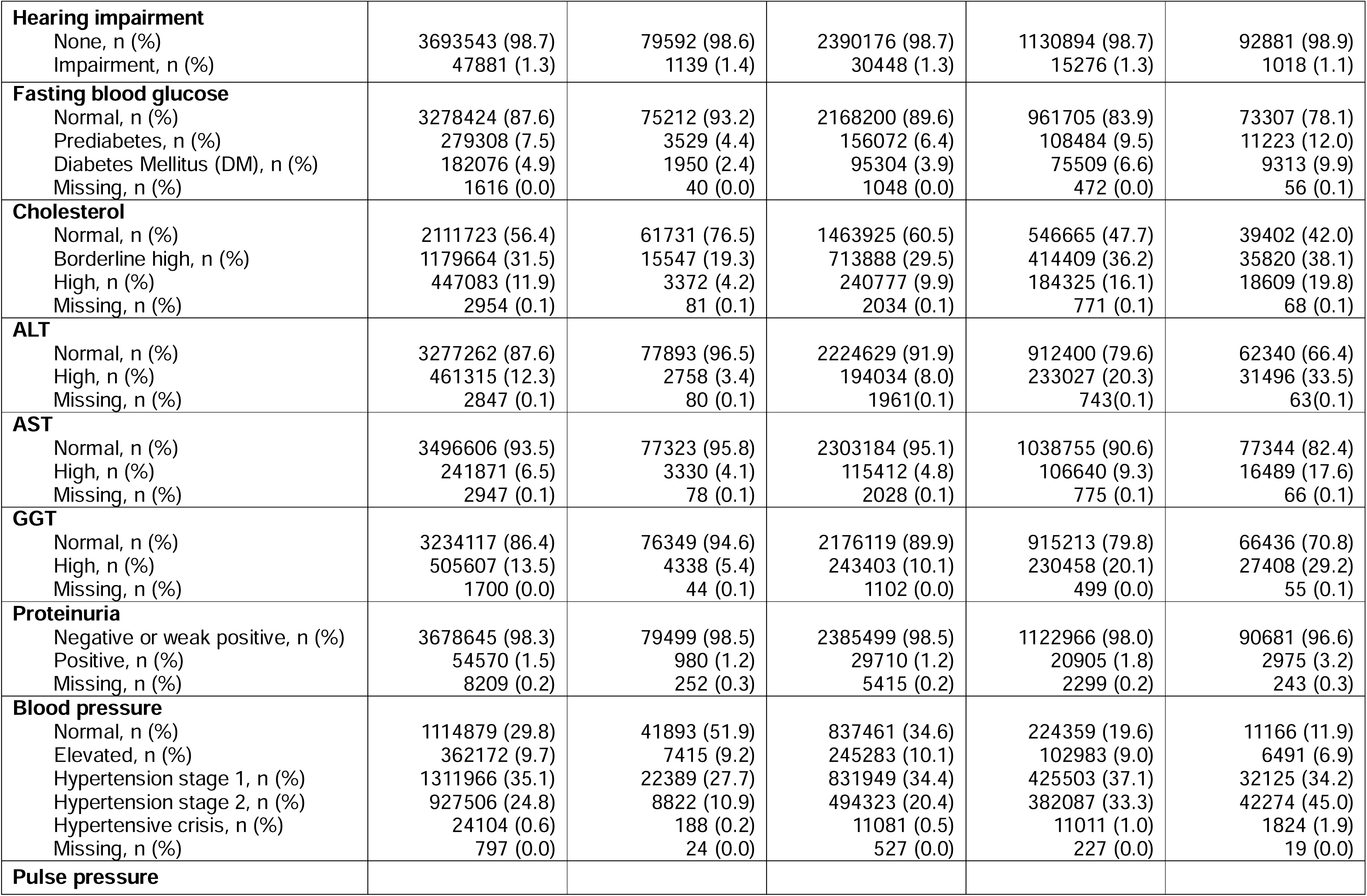
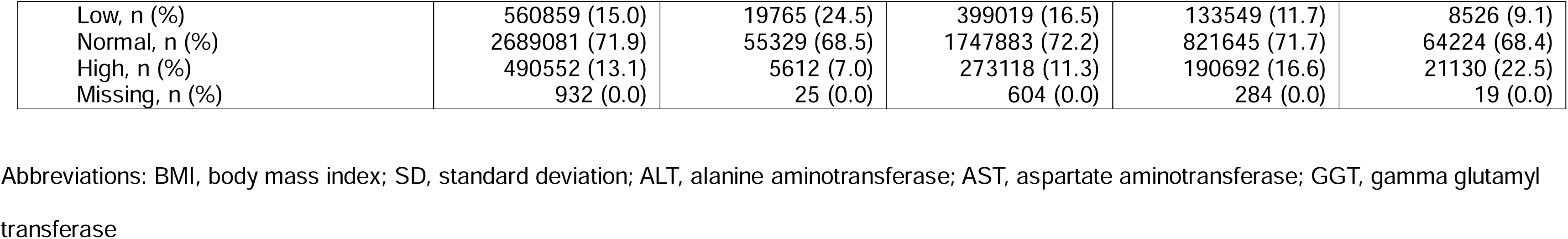
Baseline Characteristics of the Participants Stratified by the Baseline BMI.

### BMI and Development of AD

**Table 2** shows the number of events, person-years, and IRs according to the range of baseline BMI in each group classified by age and gender. IRs of underweight, overweight, and obese groups were 46.74, 44.14, and 48.88, respectively, compared to the normal weight group. When stratified by age, the oldest age group (60 to 79) had higher IRs compared to other age groups.

**Table 2.** Events, Person-years, and IRs for AD According to Baseline BMI Stratified with Gender and Age.

|  |  | Baseline BMI |  |  |  |  |  |  |  |
| --- | --- | --- | --- | --- | --- | --- | --- | --- | --- |
|  |  | <18.5 kg/m<sup>2</sup> |  |  |  | 18.5-25 kg/m<sup>2</sup> |  |  |  |
| Variable | Value | Events | Person-years | IR | 95% CI | Events | Person-years | IR | 95% CI |
| Total |  | 2086 | 446313 | 46.74 | 44.78-48.79 | 51963 | 13455512 | 38.62 | 38.29-38.95 |
| Gender | Male | 1070 | 219110 | 48.83 | 45.99-51.85 | 24097 | 7989521 | 30.16 | 29.78-30.54 |
|  | Female | 1016 | 227203 | 44.72 | 42.05-47.55 | 27866 | 5465991 | 50.98 | 50.39-51.58 |
| Age | 30-44 | 50 | 289234 | 1.73 | 1.31-2.28 | 1575 | 7052884 | 2.23 | 2.13-2.35 |
|  | 45-59 | 332 | 99076 | 33.51 | 30.09-37.32 | 13291 | 4680684 | 28.40 | 27.92-28.88 |
|  | 60-79 | 1704 | 58003 | 293.78 | 280.16-308.06 | 37097 | 1721943 | 215.44 | 213.26-217.64 |
|  |  | 25-30 kg/m<sup>2</sup> |  |  |  | ≥30 kg/m<sup>2</sup> |  |  |  |
| Variable | Value | Events | Person-years | IR | 95% CI | Events | Person-years | IR | 95% CI |
| Total |  | 27959 | 6334400 | 44.14 | 43.62-44.66 | 2448 | 500838 | 48.88 | 46.98-50.85 |
| Gender | Male | 10819 | 4218476 | 25.65 | 25.17-26.13 | 569 | 282009 | 20.18 | 18.59-21.9 |
|  | Female | 17140 | 2115924 | 81.00 | 79.80-82.23 | 1879 | 218829 | 85.87 | 82.07-89.84 |
| Age | 30-44 | 632 | 2854459 | 2.21 | 2.05-2.39 | 52 | 229989 | 2.26 | 1.72-2.97 |
|  | 45-59 | 7741 | 2501540 | 30.94 | 30.26-31.64 | 665 | 192349 | 34.57 | 32.04-37.30 |
|  | 60-79 | 19586 | 978401 | 200.18 | 197.40-203.01 | 1731 | 78500 | 220.51 | 210.36-231.15 |

Abbreviations: IR, incidence rate; AD, Alzheimer's disease; BMI, body mass index; CI, confidence interval.

**Table 3** shows the HRs for AD according to baseline BMI. Similar to the IRs, crude hazard ratios (HRs) of each of groups were as follows: underweight (HR,1.21, 95% CI, 1.16-1.27); overweight (HR,1.14, 95% CI, 1.13-1.16); and obese (HR,1.27, 95% CI, 1.22-1.32). After adjustment of comorbidities and risk factors, however, the risk for AD in the overweight (adjusted HR [aHR],1.00, 95% CI, 0.99-1.02) and obese (aHR,1.04, 95% CI, 0.99-1.08) became reduced and not statistically significant. Being underweight remained as a risk factor for AD in those 45 years of age or older (45 to 60: aHR,1.12, 95% CI, 1.00-1.25; ≥60: aHR,1.15, 95% CI, 1.10-1.21), but not in those under 45. The adjusted variables and their aHRs for AD are provided in Supplemental Figure 3.

**Table 3.**
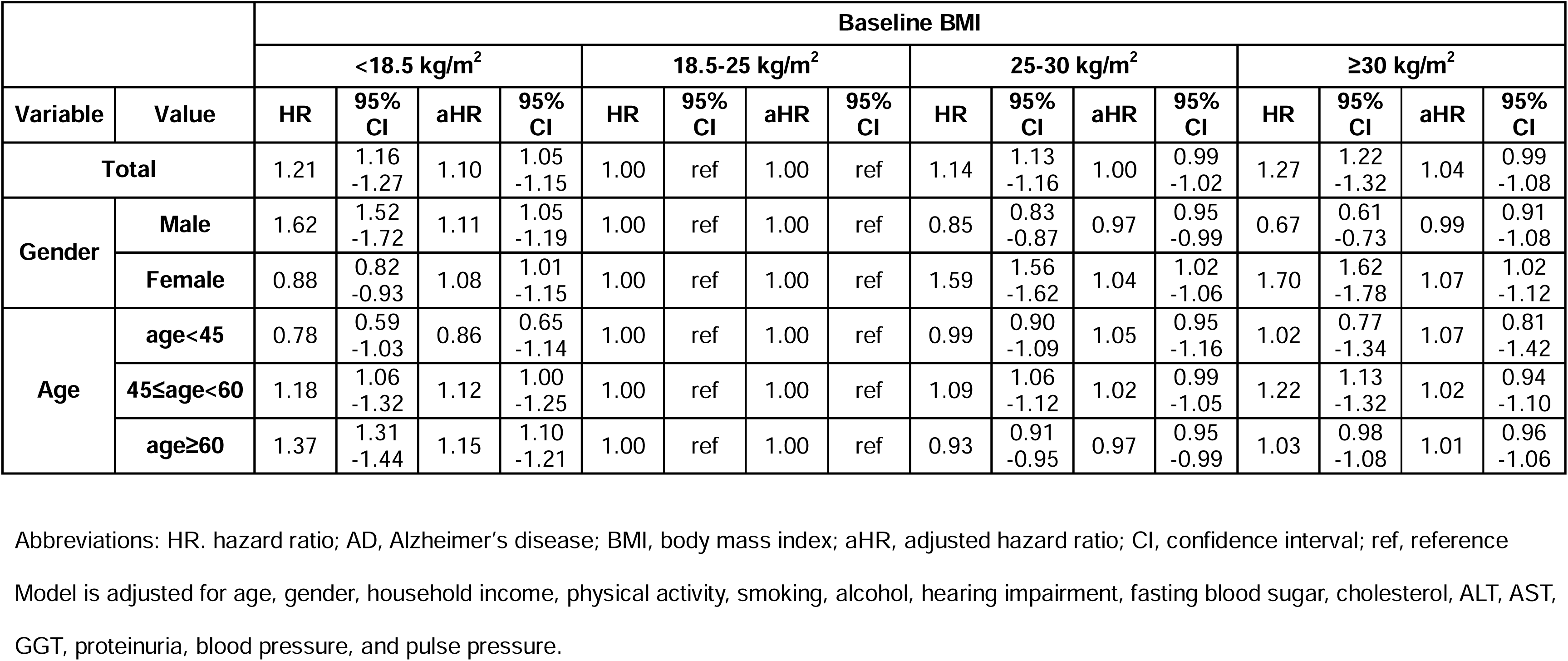
Crude and Adjusted HRs for AD According to Baseline BMI Stratified with Gender and Age.

### Impact of Weight Changes on AD Development

The risk of AD development according to the type of body weight change is shown in **Table 4**. In the underweight group, an acute increase in body weight (aHR,1.45, 95% CI, 1.04-2.01) increased the risk of AD the most but not in other groups. However, an acute decrease in body weight significantly increased the risk of AD in these groups (normal weight: aHR,2.00, 95% CI, 1.63-2.44; overweight: aHR,1.65, 95% CI, 1.32-2.07; obese: aHR,1.57, 95% CI, 1.14-2.15). Weight cycling and gradual decreases in body weight also increased the risk of AD in all BMI groups. When categorized by 3 groups of age, an acute decrease in body weight significantly increased the risk of AD in those over 60 years of age (aHR,1.92, 95% CI, 1.66-2.21). A steady decrease also increased the risk of AD regardless of age (<45: aHR,1.29, 95% CI, 1.14-1.46; 45 to 60: aHR,1.33, 95% CI, 1.28-1.38; ≥60: aHR,1.34, 95% CI, 1.31-1.37). Of note, weight cycling further increased the risk of AD (<45: aHR,1.32, 95% CI, 1.18-1.46; 45 to 60: aHR,1.43, 95% CI, 1.38-1.48; and ≥ 60: aHR,1.39, 95% CI, 1.36-1.42). An acute increase in body weight also increased the risk of AD in those aged 45 to 60 (aHR,1.39, 95% CI, 1.01-1.93).

**Table 4.** Adjusted HRs for AD According to Body Weight Changes Stratified with Baseline BMI and Age.

|  |  | Type of body weight change |  |  |  |  |  |  |  |  |  |  |  |
| --- | --- | --- | --- | --- | --- | --- | --- | --- | --- | --- | --- | --- | --- |
|  |  | Stable<br>(n = 1281954) |  | Acute Increase<br>(n = 5623) |  | Steady Increase<br>(n = 808139) |  | Weight Cycling<br>(n = 979856) |  | Acute Decrease<br>(n = 3336) |  | Steady Decrease<br>(n = 662516) |  |
| Variable | Value | aHR | 95% CI | aHR | 95% CI | aHR | 95% CI | aHR | 95% CI | aHR | 95% CI | aHR | 95% CI |
| Total |  | 1.00 | ref | 1.25 | 1.05<br>-1.48 | 1.11 | 1.09<br>-1.14 | 1.37 | 1.35<br>-1.40 | 1.78 | 1.55<br>-2.03 | 1.33 | 1.30<br>-1.35 |
| Baseline BMI<br>(kg/m<sup>2</sup>) | <18.5 | 1.00 | ref | 1.45 | 1.04<br>-2.01 | 1.21 | 1.04<br>-1.41 | 1.34 | 1.17<br>-1.53 | 0.99 | 0.14<br>-7.04 | 1.28 | 1.09<br>-1.51 |
|  | 18.5-25 | 1.00 | ref | 1.16 | 0.94<br>-1.44 | 1.12 | 1.08<br>-1.15 | 1.37 | 1.34<br>-1.41 | 2.00 | 1.63<br>-2.44 | 1.33 | 1.29<br>-1.36 |
|  | 25-30 | 1.00 | ref | 1.19 | 0.59<br>-2.37 | 1.09 | 1.05<br>-1.14 | 1.38 | 1.34<br>-1.43 | 1.65 | 1.32<br>-2.07 | 1.33 | 1.29<br>-1.37 |
|  | ≥30 | 1.00 | ref | NA | NA | 0.98 | 0.83<br>-1.15 | 1.23 | 1.10<br>-1.37 | 1.57 | 1.14<br>-2.15 | 1.30 | 1.17<br>-1.45 |
| Age<br>(year) | <45 | 1.00 | ref | 1.36 | 0.51<br>-3.65 | 1.04 | 0.93<br>-1.16 | 1.32 | 1.18<br>-1.46 | 2.13 | 0.68<br>-6.64 | 1.29 | 1.14<br>-1.46 |
|  | 45-60 | 1.00 | ref | 1.39 | 1.01<br>-1.93 | 1.15 | 1.11<br>-1.20 | 1.43 | 1.38<br>-1.48 | 1.51 | 1.01<br>-2.25 | 1.33 | 1.28<br>-1.38 |
|  | ≥60 | 1.00 | ref | 1.17 | 0.95<br>-1.44 | 1.12 | 1.09<br>-1.15 | 1.39 | 1.36<br>-1.42 | 1.92 | 1.66<br>-2.21 | 1.34 | 1.31<br>-1.37 |

Abbreviations: HR, hazard ratio; AD, Alzheimer's disease; BMI, body mass index; aHR, adjusted hazard ratio; CI, confidence interval, NA, Not applicable or not available.
Model is adjusted for age, gender, household income, physical activity, smoking, alcohol, hearing impairment, fasting blood sugar, cholesterol, ALT, AST, GGT, proteinuria, blood pressure, and pulse pressure.

### Physical Activity’s Influence on AD Risk from Weight Changes

**Figure 1** shows the risk of AD by weight changes mitigated by regular physical activities. The risk of AD development due to an acute increase (aHR,0.94, 95% CI, 0.35-2.52), steady increase (aHR,1.04, 95% CI, 0.94-1.15), and acute decrease (aHR,1.19, 95% CI, 0.45-3.21) disappeared in participants with regular physical activity. However, in participants who did not regularly engage in physical activity, all weight changes remained significant risks for AD.

**Figure 1.**
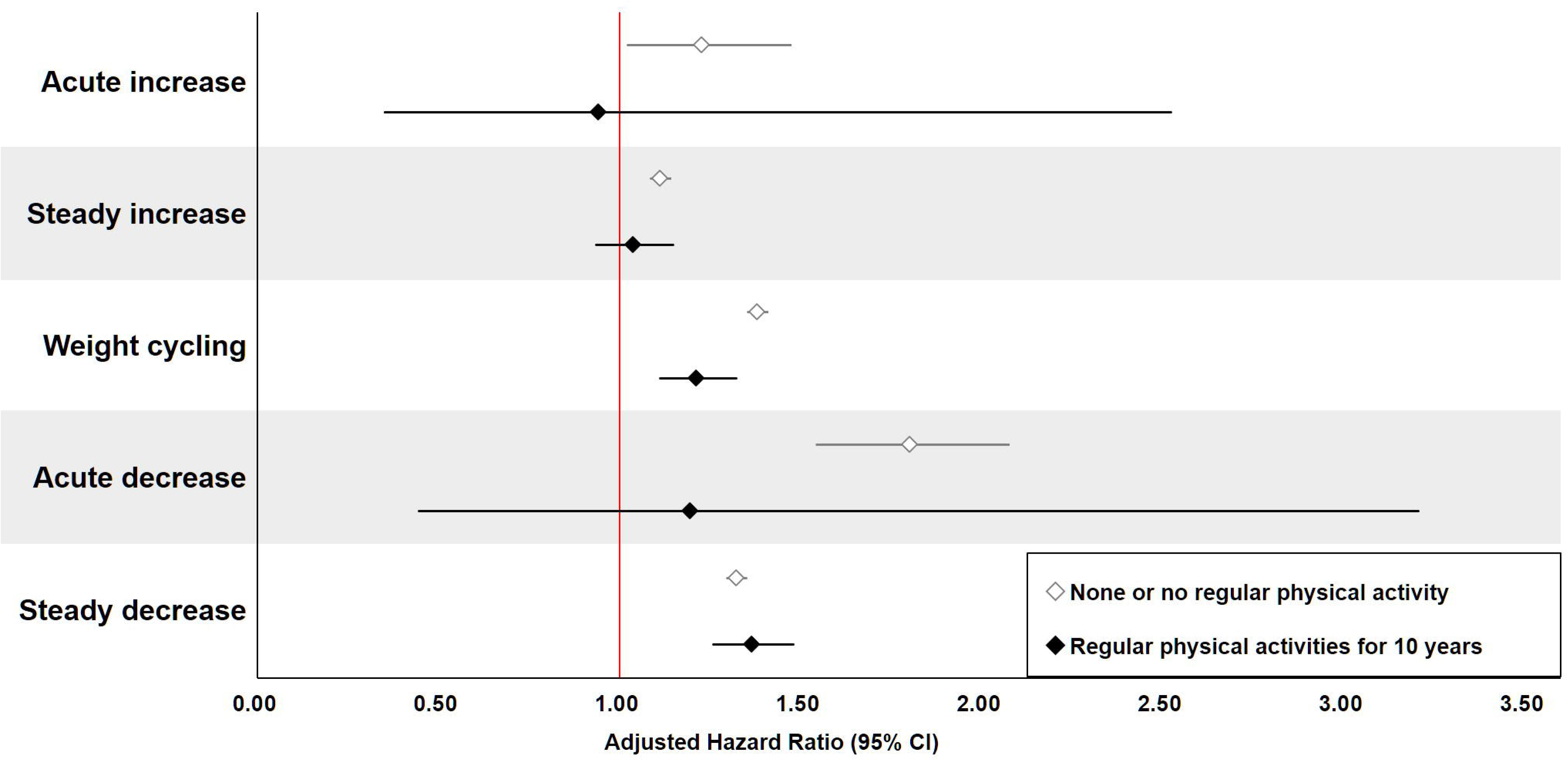
Adjusted HRs for AD by Body Weight Changes, Stratified by Regular Physical Activities for a Decade. The individuals with regular physical activity are those who have performed at least one physical activity per week for 10 years. The group with no or irregular physical activity consists of those who do not exercise or exercise irregularly, not fulfilling the definition of regular physical activity. The reference HR (1.00) is indicated by a red line.

## Discussion

This study examined the impact of body weight on AD development. In contrast to other studies that focused on BMI at a single time point or short-term BMI changes, this is the first to analyze weight changes over a 10-year period with six biennial assessments, including detailed health metrics and laboratory data. Unlike prior findings, BMI alone did not affect AD development after adjusting for risk factors. Weight change, however, was an obvious risk factor for AD. The effect of weight change on AD development was consistent across most groups, although it varied by age or baseline BMI. Regular physical activity reduced the risk of AD related to weight changes.

### Does BMI Affect the Development of AD?

In previous studies, the effect of BMI on the development of AD varied by age.^22^ During middle age, a high BMI was identified as a risk factor for AD, whereas in older age, a low BMI increased the risk of AD and high BMI worked as protective factor for AD, which is a phenomenon also known as the obesity paradox.^11^ Weight loss after AD onset further complicates the body weight-AD association.^18^ ^19^ In our study, being underweight, overweight, and obese all heightened the risk of AD when other variables were not considered. However, after adjusting for known risk factors for AD, baseline BMI values no longer increase the risk of AD, except for being underweight in seniors (aged ≥60 years), which retained its risk for AD. Our findings suggest that midlife obesity is not a risk factor for AD merely as a simple measurement of body weight, but it may affect AD development by influencing other factors such as socioeconomic status and DM (**Supplemental Figure 3**) in line with previous research findings.^23^

### Impact of Weight Changes on AD Development

The most notable findings were the increased risk of AD in the acute increase and acute decrease groups. Particularly, an acute decrease had a stronger effect on AD development than other body weight changes in subjects with normal baseline BMI. Conversely, an acute increase in the underweight group also showed detrimental effects on AD development. These findings suggest that rapid weight correction may exacerbate AD development, indicating the need to consider the effect of weight changes prior to correction. Additionally, both acute and gradual weight decreases increased AD risk compared to stable body weight. These results align with studies demonstrating that adequate nutritional intake can be a preventive factor for AD^34^ and that nutritional deficiencies worsen AD progression.^24^ ^25^

Weight cycling had overall negative effects on AD development. Repetitive weight loss can lead to muscle loss, reduced metabolism and physical strength.^26^ Thus, each cycle of weight loss and gain may slow the metabolic rate and complicate future weight loss efforts.^27^ Moreover, when weight is regained, it often returns as fat, not muscle, potentially worsening body composition^28^ and causing hormonal disturbances related to appetite, nutritional metabolism, and energy homeostasis, which contribute to AD pathogenesis.^8^

### Impact of Weight Changes on AD development Stratified by Age

Also, the effect of weight change on AD varied by age. Weight loss, weight cycling, and weight gain all increased the risk of AD in older groups (≥45), but only weight loss and weight cycling were risk factors for AD in the youngest group (30-44). It is notable that the weight gain-associated AD risk was only noted in those over 45 years old. Similar age- and weight change-associated findings have been reported regarding mortality in healthy individuals (e.g. the higher mortality associated with weight gain in the elderly).^29^ Given that muscle mass decreases and becomes difficult to regain as people get older,^30^ ^31^ it is plausible that weight gain in those over 45 years is most likely to be obesity that reduces muscle mass or myosteatosis,^32^ contributing to the study results.

Additionally, it has been reported in the elderly that weight change accompanied by weight loss is a risk factor for AD.^29^ Our findings demonstrated that weight loss can also be a risk factor for AD even in younger individuals (<45). Both findings may contribute to brain changes associated with intended or unintended nutritional deficits, decrease in muscle mass, deteriorated immune function, and underlying diseases.^29^ ^33^

### Physical Activity Mitigating AD Risk Related to Weight Changes

The strong association between weight changes and AD development was attenuated with regular exercise for a decade. The most prominent effect of physical activity on reducing the risk of AD was observed in subjects with either an acute increase or acute decrease in body weight, which implicates the potential role of consistent exercise in muscle gain or fat loss, respectively. Multifactorial benefits of regular exercise such as increased cerebral blood flow, enhanced neuroplasticity, release of neurotrophic factors, decreased inflammation, and increased insulin resistance may have contributed to the mitigation.^34^ Increased brain metabolism and better clearance of cerebral waste products have also been suggested as beneficial effects of regular exercise.^35^

### Limitations and Strength of the Study

This study has a couple of limitations. First, AD diagnosis relied solely on diagnostic codes without consideration for biomarkers for assessing the AD pathological changes. However, the authors attempted to investigate most of the available risk factors for AD as shown in **Table 1**. In addition, a recent research comparing diagnostic accuracy between diagnostic codes alone and both diagnostic codes and medication history has shown that the combined approach often underestimates the actual prevalence of AD.^36^ In contrast, using only diagnostic codes closely mirrors the true prevalence of AD, which is the approach we have applied in this study to define the occurrence of AD.^36^

Second, Korea has the highest proportion of people with body weight in the normal ranges in the world.^7^ Over 60% of the Korean population falls into the normal BMI category, indicating a scarcity of underweight and obese individuals. Among the obese, the proportion of individuals with a class I obesity (30≤BMI<35) in Korea is about 90%,^7^ which aligns closely with our findings (class I obesity: 93.7%; class II obesity (35 ≤BMI<40): 5.0%; class III obesity (BMI>40): 1.3%). This higher proportion of class I obesity contrasts sharply with the global average of about 70% and only 50-60% in the United States (**Supplemental Figure 4, Supplemental Method 4**).^7^ The lower prevalence of severe obesity in Korea, particularly class III obesity, may weaken the association between obesity and AD, potentially due to a reduced prevalence of obesity-associated metabolic changes, such as insulin resistance, which is an important factor in AD development.^37^

Third, considering the significantly increased risk of AD associated with weight loss, it becomes imperative to differentiate the impacts of intended versus unintended weight loss. In this community-based cohort study, information regarding the participants’ diets or their intentions for weight control were not obtained. To address this limitation, we utilized comprehensive physical activity records as a proxy to gauge the participants’ commitment to intentional weight management. Consistent with the observed protective effect of physical activity against AD risk, it is plausible that individuals who engage in regular and intensive physical activity—a likely marker of active weight management—could have reduced their AD risk.^38^ ^39^

With nearly 100% coverage by the Korean NHIS, the study findings can be representative of the entire population of the Republic of Korea. Data reliability with minimal missing entries is ensured through objective and detailed measurements and recordings by authorized medical institutions.^40^ Additionally, we tried to remove the potential effect of weight loss that can occur after the development of AD, by adding a 1-year post-washout period after the decade of body weight measurement, focusing on weight changes preceding AD development.

## Conclusion

Changes in body weight, instead of BMI alone, are significant risk factors for the development of AD. Regular physical activity mitigated these risks. Public health recommendations should emphasize the importance of maintaining a stable weight and encouraging consistent physical activity, rather than focusing solely on preventing obesity and weight loss during mid-to-late adulthood.

## Acknowledgments

The authors appreciate the support from the Myokine Convergence Research Center.

## Contributors

RJH contributed to conceptualization, funding acquisition, investigation, methodology, resources, supervision, visualization, and writing original draft. JI contributed to conceptualization, data curation, investigation, methodology, visualization, and writing original draft. KHJ contributed to conceptualization, data curation, investigation, methodology, resources, supervision, and writing original draft.

## Patient consent for publication

Not applicable.

## Data availability statement

Data are available upon reasonable request.

