## Supplemental for "Increased Risk of Alzheimer’s Disease Affected by Weight Changes but Not by Body Mass Index"

**Supplemental Figure 1.** Study Flow Diagram
**Supplemental Figure 2.** Study Design
**Supplemental Figure 3.** Adjusted Hazard Ratio (aHR) for Alzheimer's Disease Associated with Variables Shown in Table 1
**Supplemental Figure 4.** A Classification of Obese Population by 3 BMI Units in Korea, Worldwide and the USA (2003-2016)

**Supplemental Method 1.** Data Sources
**Supplemental Method 2.** Comorbidities or Risk Factors
**Supplemental Method 3.** The Composition of the Obese Population Worldwide

**Supplemental Figure 1. Study Flow Diagram**

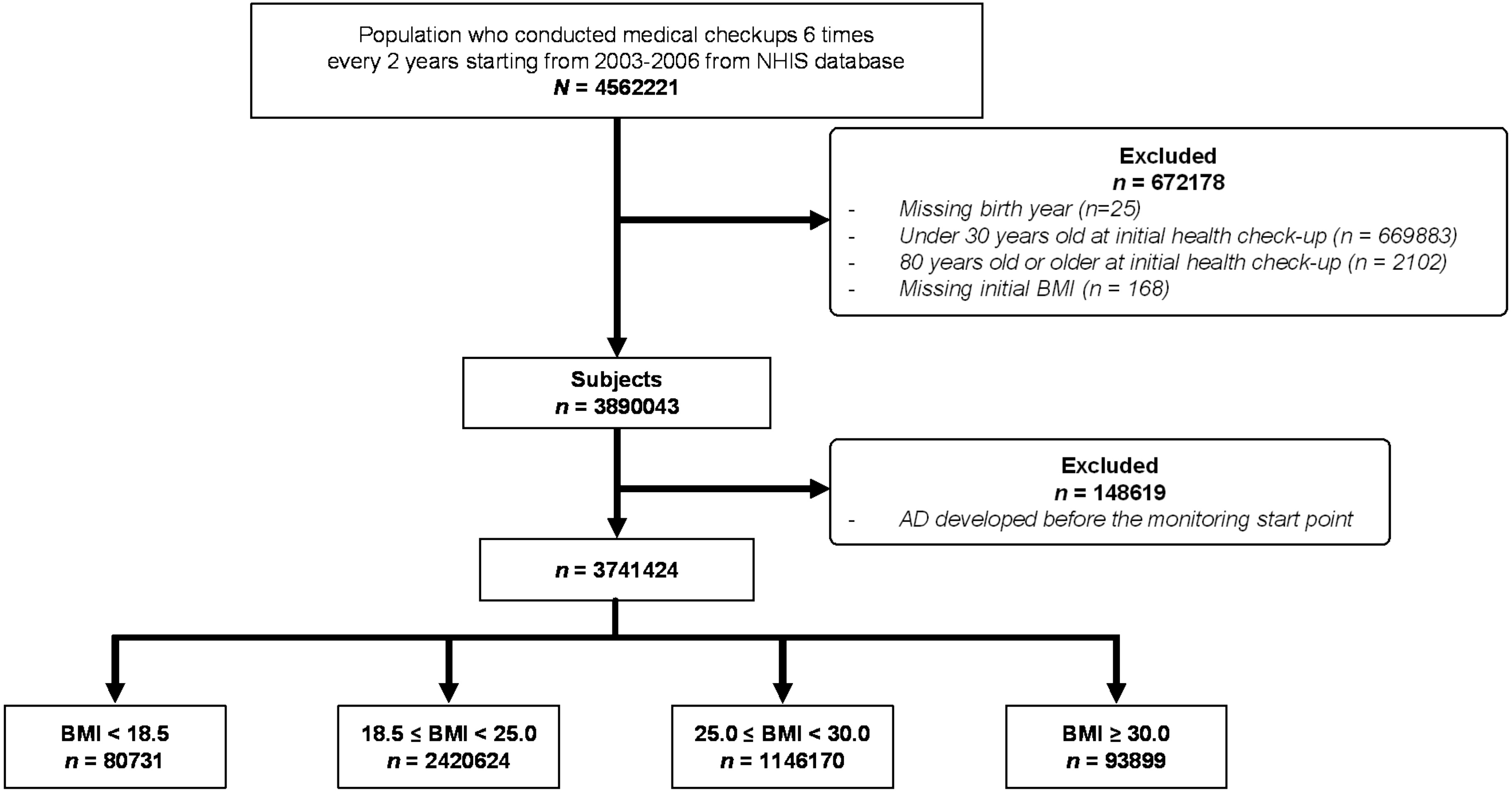


This flow diagram illustrates the participant selection process. The unit for BMI was standardized to $kg/m^2$.

### Supplemental Figure 2. Study Design

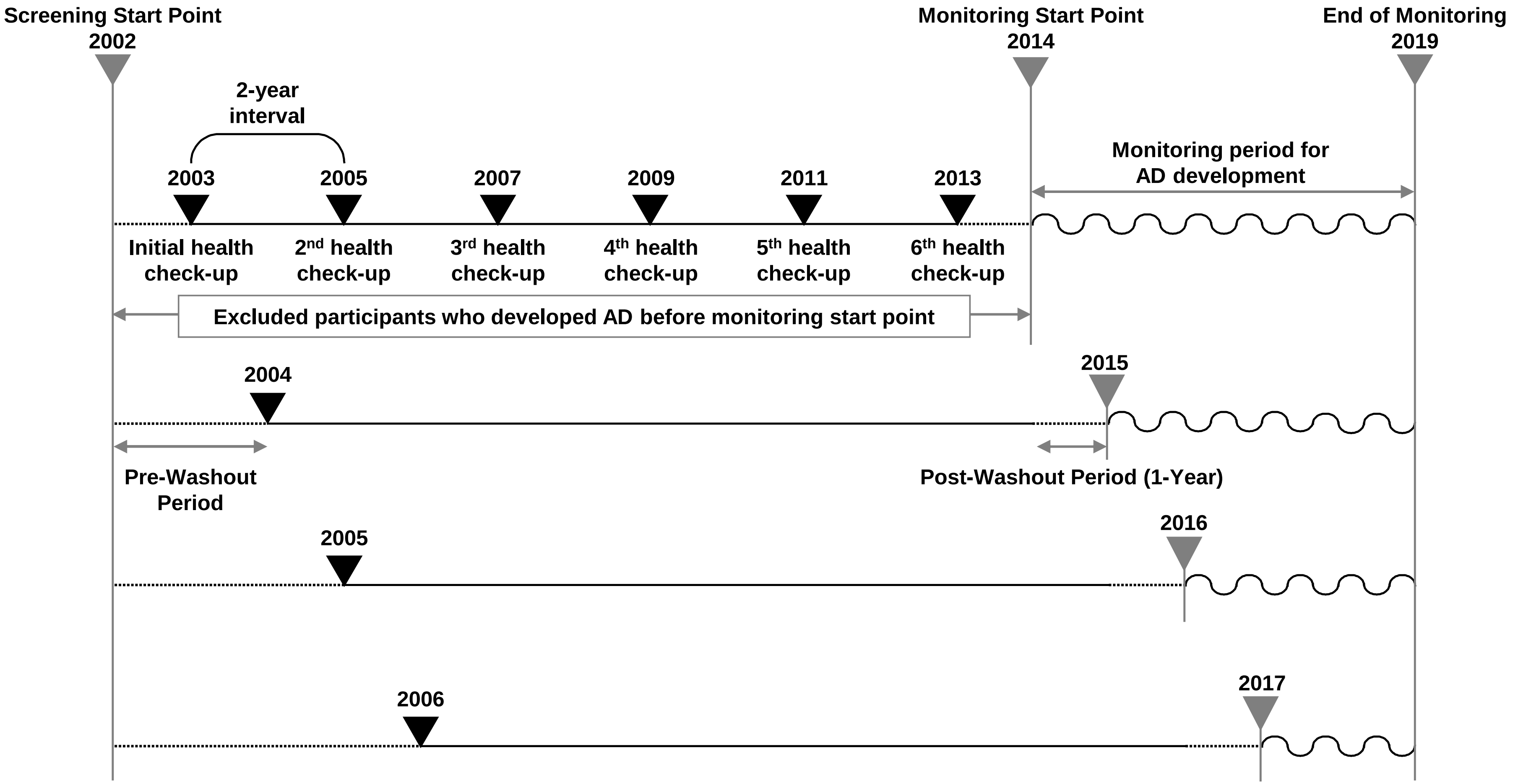


Biennial health check-ups were conducted six times over ten years, began in 2003, 2004, 2005, and 2006, each following a pre-washout period. Monitoring for Alzheimer's Disease (AD) commenced after an additional one-year post-washout period for each group. Participants who developed AD before their respective monitoring start points were excluded. Dashed lines indicate the washout periods before and after the health evaluation periods and curved lines denote the monitoring periods for AD development.

### Supplemental Figure 3. Adjusted Hazard Ratio (aHR) for Alzheimer's Disease Associated with Variables Shown in Table 1

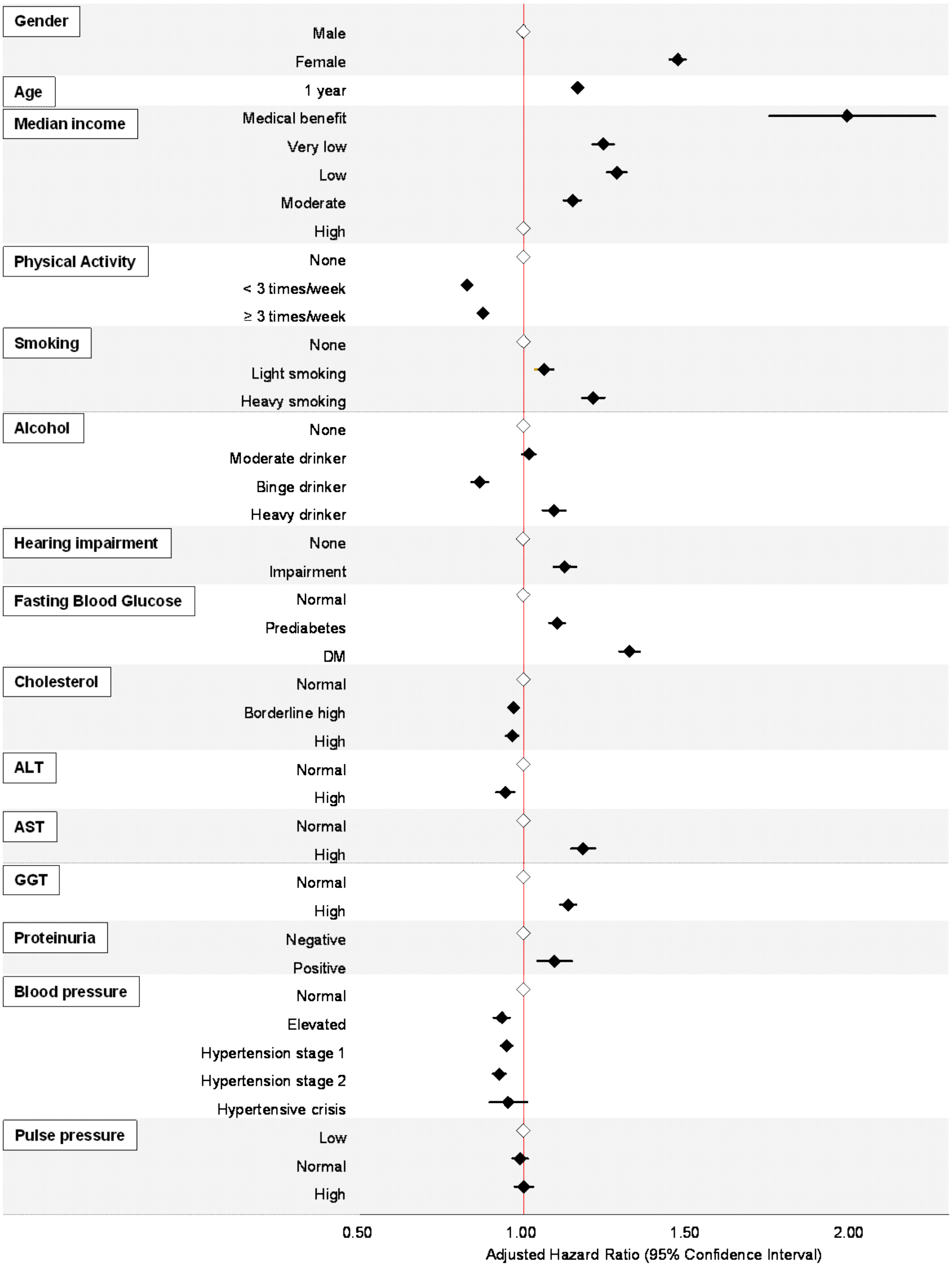


The model is adjusted for baseline BMI and all variables except for the one being analyzed, as shown in the left column. Detailed variable categories and their aHRs are provided. The reference HR (1.00) is indicated by a red line. A blank diamond represents the reference for each variable, the definition of which is provided in **Supplemental Method 2**.

### Supplemental Figure 4. A Classification of Obese Population by 3 BMI Units in Korea, Worldwide and the USA (2003-2016)

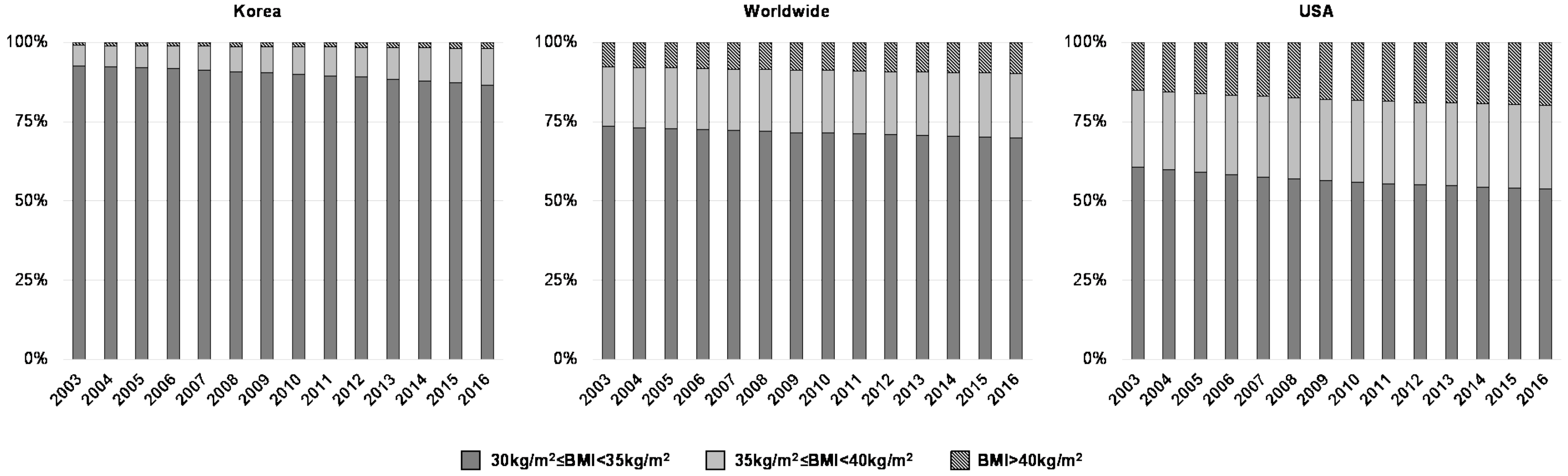


The proportions of Class I-III obesity were recalculated using data from 'Worldwide trends in underweight and obesity from 1990 to 2022,' *The Lancet* 2024 paper.[1] The period from 2003 to 2016 was selected to correspond to the baseline body weight assessment in this study. Detailed methods for calculation are provided in **Supplemental Method 3**.

### **Supplemental Method 1.** Data Sources

**National Health Insurance (NHI)**
The NHI is a Korean government-operated service that provides mandatory insurance to the nation's entire population.[2] Employed or self-employed individuals become NHI beneficiaries by paying an insurance premium corresponding to their income. The NHI includes both inpatient and outpatient healthcare utilization data, including prescriptions, procedures and diagnoses classified according to the International Classification of Disease, 10th revision (ICD-10).[3] The NHI database also includes demographic information such as age, sex, employment status and insurance premiums. NHI data were collected from 2002 to 2019.

**National Health Screening Program (NHSP)**
The NHSP is a screening program that offers biennial standardized health check-ups to all NHI beneficiaries, the results of which are recorded in the database.[4] The examination includes a standardized questionnaire on participants' medical history and lifestyle habits, including physical activity, smoking and alcohol consumption. Anthropometric measurements and basic tests are also taken at the check-up, including BMI, systolic and diastolic blood pressure, blood tests, urine tests, and chest radiographs.

**National Disability Registration System (NDRS)**
The NDRS is a social welfare program that provides benefits to individuals with disabilities.[5] In order to be registered in the program, the individual must be diagnosed by a qualified physician according to standardized diagnostic criteria. The degree of disability is also assessed. Information on hearing loss was acquired through the NDRS.

### **Supplemental Method 2.** Comorbidities or Risk Factors

Smoking was categorized according to pack-years. We defined men who drank less than 5 units and women who drank less than 4 units per drink as "moderate drinkers." Men who drank more than 5 units and women who drank more than 4 units per drink were classified as a "binge drinker" if they drank less than once a week, and a "heavy drinker" if they drank more than once a week.[6]

Fasting blood glucose was classified as normal (<110 mg/dL), prediabetes (≥110 mg/dL and <126 mg/dL), and diabetes mellitus (DM ≥126 mg/dL).[7] Cholesterol levels were divided into normal (<200 mg/dL), borderline high (200 mg/dL or more and less than 240 mg/dL), and high (≥240 mg/dL).[8] ALT and AST were classified as normal (≤40 mU/mL) and high (>40 mU/mL). GGT levels were classified as normal (men <63 mU/mL; women <35 mU/mL) and high (men ≥63 mU/mL; women ≥35 mU/mL). Proteinuria was classified into a negative or weakly positive group and a positive group.

Blood pressure was classified as normal, elevated, hypertension stage 1, hypertension stage 2, and hypertensive crisis.[9] Pulse pressure was classified as low (<40 mmHg), normal (≥40 mmHg and <60 mmHg), and high (≥60 mmHg).

The insurance premiums paid by beneficiaries were used as a proxy for socioeconomic status, as they are calculated based on an employee's wage. We categorized the study population into 5 groups according to income level, with the first group including individuals who are medical benefit recipients (those under the poverty line or with no stable source of income).

### **Supplemental Method 3.** Three different Cox proportional hazard regression models

**Effect of BMI on AD development**
To examine the association between BMI and AD, stratified Cox proportional hazard regression models were used and hazard ratios (HR) were calculated with 95% confidence intervals (CIs). HRs compared the risk of AD among underweight, overweight and obese individuals with normal weight individuals (reference group). HRs were adjusted for risk factors, including age, sex, income level, physical activity, smoking, alcohol consumption, hearing impairment, fasting blood glucose, cholesterol, ALT, AST, GGT, proteinuria, blood pressure, and pulse pressure. The proportional hazard assumption was tested by using the Schoenfeld assumption and scale Schoenfeld residuals. Separate age- and sex-stratified analyses were also conducted.

**Effect of body weight changes on AD development**

In another Cox model, we evaluated the effect of body weight change on AD. HRs assessing the risk of AD among individuals with body weight change in 5 categories, with stable weight individuals as the reference group, were measured and adjusted for risk factors. Because the effect of body weight change on AD may vary according to one's baseline BMI and age, we further stratified the groups for 4 BMI categories and three age groups (30 to 44, 45 to 59, and 60 to 79 years).

**Effect of physical activity on AD development**

The effect of regular physical activity was assessed in another Cox model. The adjusted HRs for AD according to the type of weight change were stratified according to the status of physical activity over a 10-year period (no regular physical activity or regular physical activity) with the same adjustment of comorbidities and risk factors except the addition of regular physical activity We used Stata version 15.0 (Stata Corp) in the execution of all statistical analyses.

### **Supplemental Method 4.** The Composition of the Obese Population Worldwide

To better understand the composition of the obese population in our study and to verify the proportions of class I-III obesity among our participants, we calculated the composition of the obese populations of Korea, the world, and the United States using data provided in a recent publication.[1] Data were collected and analyzed using the NCD Risk Factor Collaboration (NCD-RisC) from 2003 to 2016, the period during which our study participants received their health check-ups. The obese population was classified into three groups: class I obesity (30≤BMI<35), class II obesity (35≤BMI<40), and class III obesity (BMI≥40). **Supplemental Figure 4** shows the percentage distribution of the obese population in each group during the study's screening period.

### Supplemental Content References

1. Collaboration NCDRF. Worldwide trends in underweight and obesity from 1990 to 2022: a pooled analysis of 3663 population-representative studies with 222 million children, adolescents, and adults. *Lancet.* 2024;403(10431):1027-1050.
2. Lee J, Lee JS, Park S-H, Shin SA, Kim K. Cohort profile: the national health insurance service–national sample cohort (NHIS-NSC), South Korea. *International journal of epidemiology.* 2017;46(2):e15-e15.
3. World Health Organization. International Statistical Classification of Diseases and Related Health Problems, 10th Revision (ICD-10). 2019; https://icd.who.int/browse10/2019/en. Accessed 18.Aug.2024.
4. Shin DW, Cho J, Park JH, Cho B. National General Health Screening Program in Korea: history, current status, and future direction. *Precision and Future Medicine.* 2022;6(1):9-31.
5. Kim M, Jung W, Kim SY, Park JH, Shin DW. The Korea national disability registration system. *Epidemiology and health.* 2023;45.
6. National Institute on Alcohol Abuse and Alcoholism. Drinking Levels and Patterns Defined. 2024; https://www.niaaa.nih.gov/alcohol-health/overview-alcohol-consumption/moderate-binge-drinking. Accessed 18.Aug.2024.
7. American Diabetes Association. Understanding Diabetic Diagnosis (Blood Glucose & AC1). 2024; https://diabetes.org/about-diabetes/diagnosis. Accessed 18.Aug.2024.
8. White H, Pieper C, Schmader K. The association of weight change in Alzheimer's Disease with severity of disease and mortality: A longitudinal analysis. *J Am Geriatr Soc.* 1998;46(10):1223-1227.
9. American Heart Association. Understanding Blood Pressure Readings. 2024; https://www.heart.org/en/health-topics/high-blood-pressure/understanding-blood-pressure-readings. Accessed 18.Aug.2024.
